# Integrated Neuronal Injury and Dysregulated Wnt Signaling Are Associated with Chronic Fatigue Syndrome and Psychiatric Symptoms in Parkinson’s Disease

**DOI:** 10.64898/2026.03.15.26348456

**Authors:** Tabarek Hadi Al-Naqeeb, Hussein Kadhem Al-Hakeim, Yingqian Zhang, Michael Maes

**Author notes:** Corresponding Author: Prof. Michael Maes, M.D., Ph.D. International NIMETOX Center, Sichuan Provincial Center for Mental Health Sichuan Provincial People’s Hospital, School of Medicine, University of Electronic Science and Technology of China, Chengdu 610072, China.

## Abstract

**Background:** Parkinson’s disease (PD) is a progressive neurodegenerative disorder with complex pathophysiology. The potential of integrating biomarkers of neuronal injury, neuroinflammation, and modulators of Wnt signaling for PD diagnosis remains largely unexplored.

**Objective:** This study aimed to evaluate the diagnostic and clinical predictive value of a ten-biomarker serum panel encompassing markers of neuronal injury (NSE, UCHL1), neuroinflammation (GFAP, HMGB1), synaptic plasticity (BDNF), proteinopathy (α-Synuclein, β-Amyloid-42), and Wnt signaling (R-spondin-1, DKK1, Sclerostin), with a particular focus on chronic fatigue in PD.

**Methods:** In this case-control study, 90 PD patients and 45 healthy controls were enrolled. Serum biomarkers were quantified using ELISA. Clinical severity was assessed using the Movement Disorder Society-Unified Parkinson’s Disease Rating Scale (MDS-UPDRS) and Fibro-Fatigue (FF) scales. Binary logistic regression and multiple linear regression analyses were used to evaluate the diagnostic and predictive value of biomarkers for PD diagnosis, psychiatric and motoric scores, and an FF score reflecting chronic fatigue syndrome (CFS) severity.

**Results:** A model incorporating NSE, DKK1, and β-Amyloid-42 effectively discriminated PD patients from controls, yielding an area under the curve (AUC) of 0.932 and an overall accuracy of 83.0%. NSE and DKK1 emerged as the main predictors of overall disease severity, motor symptoms, and CFS severity. Regression analyses indicated that 41.3% of the variance in the FF score was explained by increased NSE, DKK1, β-amyloid, and UCHL1, while 42.9% of the variance in psychiatric symptoms was explained by increased NSE, DKK1, and β-amyloid. Increased GFAP levels were significantly associated with motor dysfunction.

**Conclusion:** The combined presence of neuronal injury, Wnt signaling dysregulation, and amyloid pathology may represent a key pathophysiological component underlying PD, CFS-like fatigue, and psychiatric symptoms in PD. Targeting neuronal injury and Wnt signaling pathways may offer novel therapeutic strategies for managing fatigue and psychiatric manifestations in PD.

## Introduction

Parkinson’s disease (PD) is the second most prevalent progressive neurodegenerative disorder, characterized by a spectrum of motor and non-motor symptoms that contribute to marked disability and impaired quality of life [1]. The core motor symptoms of PD are well described, especially bradykinesia, stiffness, tremor, and postural instability. In contrast, non-motor symptoms, such as cognitive impairment, sleep disturbances, and chronic fatigue syndrome (CFS), are increasingly acknowledged as substantial contributors to disease burden, especially in advanced disease stages [2, 3]. Among these, fatigue represents a highly debilitating yet poorly understood symptom, underscoring the urgent need for deeper insights into its underlying pathophysiology. The pathophysiology of PD involves the progressive degeneration of dopaminergic neurons in the substantia nigra, driven by complex mechanisms that include protein aggregation, neuroinflammation, and the dysregulation of cell death pathways such as apoptosis, necroptosis, and pyroptosis [4]. The formation of Lewy bodies derived from α-synuclein constitutes a pathogenic hallmark of PD, and the disruption of its inherent neuroprotective function leads to synaptic dysfunction and increased neuronal vulnerability [4–6]. Given this multifaceted complexity, identifying accurate and reliable biomarkers is critical to clarify the etiology of specific symptoms such as fatigue and to guide the development of targeted therapeutic strategies.

Studies investigating biomarkers in PD have predominantly focused on proteins reflecting core essential pathological processes, which can be broadly classified into indicators of neuronal damage, neuroinflammation, and synaptic dysfunction. Neuron-Specific Enolase (NSE) and Ubiquitin C-terminal Hydrolase L1 (UCHL1) are well-established biomarkers of neuronal damage. NSE is released by damaged neurons and can promote neuroinflammation by activating microglia [7, 8]. UCHL1 plays a crucial role in protein degradation via the ubiquitin-proteasome system; mutations and post-translational modifications of UCHL1 are implicated in PD pathogenesis by modulating α-synuclein aggregation and neuronal viability [9, 10]. Proteins involved in aggregation and neuroprotection, such as α-Synuclein and β-Amyloid-42, are crucial in the proteinopathy underlying neurodegeneration. In patients with PD, elevated serum and plasma levels of α-synuclein have been correlated with disease-related pathology [11, 12]. Alterations in β-Amyloid-42, a protein classically linked to Alzheimer’s disease, are also observed in PD and correlate with cognitive deficits, implying shared pathogenic mechanisms across neurodegenerative disorders [13, 14].

Brain-derived neurotrophic factor (BDNF) plays a crucial role in neuronal survival and synaptic plasticity. Its levels are often altered in PD and have been associated the severity of both motor and non-motor symptoms, although its role may be complex and partly compensatory [15, 16]. Glial Fibrillary Acidic Protein (GFAP) and High Mobility Group Box 1 (HMGB1) are established biomarkers for astrocytic activation and proinflammatory signaling, respectively, underscoring the importance of neuroinflammation in PD [16, 17]. Before these well-characterized markers, proteins involved in the Wnt signaling pathway-an essential regulator of brain development and synaptic preservation-represents a promising but unexplored area for biomarker research. R-spondin-1 (RSPO1), Dickkopf-1 (DKK1), and sclerostin are key modulators of this pathway [18]. However, their roles in PD remain poorly understood, and their potential to reflect synaptic dysfunction and disease progression has yet to be clarified. Moreover, few studies have investigated comprehensive biomarker panels that integrate conventional indicators of brain damage with innovative Wnt pathway modulators to predict specific, severe non-motor symptoms, such as fatigue, and overall disease severity.

Furthermore, the diagnostic performance of this multi-biomarker panel in distinguishing patients with PD from healthy controls using accessible blood samples requires further validation. This study seeks to investigate the serum concentrations of ten biomarkers-NSE, UCHL1, BDNF, α-Synuclein, β-Amyloid-42, GFAP, HMGB1, RSPO1, DKK1, and sclerostin- and to evaluate their combined utility in predicting CFS and non-motor symptom severity in PD, as well as their ability to differentiate between PD patients and healthy controls. By integrating markers of neuronal injury, neuroinflammation, and Wnt signaling, we aim to establish a more comprehensive serological profile of PD and its symptom profiles, which may improve clinical stratification and provide new insights into underlying pathophysiological mechanisms.

## Subjects and methods

### Subjects

The present case-control study included 90 patients with PD and 45 healthy controls. The specimens were collected from Al-Sadr Medical City, Al-Najaf Teaching Hospital, and Al-Furat Al-Awsat Center for Neurosciences. The patients’ assessment was conducted by taking a comprehensive medical history and performing a thorough clinical examination.

The diagnosis of PD was made according to the UK Parkinson’s Disease Society Brain Bank Clinical Diagnostic Criteria [19]. All patients exhibited bradykinesia, along with at least one other cardinal symptom (resting tremor, stiffness, or postural instability), and showed no characteristics indicative of an alternative Parkinsonian syndrome [19]. For the motor assessment, we used Part III (Motor Examination) of the MDS-UPDRS to rate MS severity [20]. To maintain consistency, patients were assessed in the ‘OFF’ drug state, after a minimum 12-hour cessation of all dopaminergic medicines. A combination of tools was used to describe the severity and burden of non-MS. The NMS scale was used for a thorough assessment [21]. The Hoehn and Yahr staging method was used to assess PD severity [22]. The same qualified neurologist gave all clinician-rated scales. The neurologist examined systemic diseases that may affect the studied parameters, specifically liver disease and kidney disease, which were excluded from the study. To rule out overt systemic inflammation, serum C-reactive protein (CRP) levels in all samples were low, with values below 6 mg/L [23]. This test was conducted to exclude the presence of overt infection or inflammation that may significantly elevate acute-phase reactant proteins, especially CRP [23].

The control and patient participants were required to give written consent before participating in the study. They were provided with detailed information beforehand. The study was granted approval by the Institutional Ethics Committee of the University of Kufa (MEC-110/2025). The study adhered to both local and international ethical and privacy laws. It complied with various international guidelines and declarations, such as the World Medical Association’s Declaration of Helsinki.

### Clinical measurements

An experienced neurologist conducted a semi-structured interview to evaluate and gather socio-demographic and clinical information from control subjects and patients. An expert in neurology assessed the extent of motor and MS symptoms associated with PD by employing the Movement Disorders Society Revision of the Unified Parkinson’s Disease Rating Scale (MDS-UPDRS), as previously outlined [24, 25]. The MDS-UPDRS is divided into four parts: Part I: non-motor Experiences of Daily Living (nM-EDL), Part II: motor Experiences of Daily Living (m-EDL), Part III: motor examination, and Part IV: motor complications [25]. Neurologists conducted the MDS-UPDRS assessment of patients, utilizing the ratings from the four domains for statistical analysis. The overall severity of PD was conceptualized as the first principal component score derived from the total nM-EDL, total m-EDL, Part III, Part IV, and Hoehn and Yahr stage scores. Psychiatric symptoms were conceptualized as the sum of the following nM-EDL items: cognitive impairment, hallucinations, depressed mood, anxious mood, apathy, sleep problems, and fatigue. The severity of CFS and fibromyalgia was assessed by a senior psychiatrist using the Fibro-Fatigue scale [26]. Consequently, we computed a “pure” fatigue score by deleting all non-CFS-related symptoms, namely irritability, sadness, insomnia, and retaining muscle pain and tension, fatigue, concentration-memory disorders, autonomous and gastrointestinal symptoms, headache, and a flu-like malaise.

### Assays

A volume of 5 milliliters of fasting blood samples was collected at approximately 9:00 a.m. After complete clotting, the blood samples were centrifuged at 1200 × g for five minutes at room temperature. Subsequently, the serum was carefully divided into three Eppendorf tubes. Excluded from the study were samples that had undergone hemolysis. The tubes were subsequently frozen at -80 °C and remained frozen until thawed for the assays.

We utilized sandwich ELISA to quantify serum levels of human NSE, UCHL1, BDNF, α-Synuclein, β-Amyloid-42, GFAP, HMGB1, RSPO1, DKK1, and sclerostin. These measurements were conducted using pre-made ELISA kits provided by Wuhan USCN Business Co., Ltd. (China). The coefficient of variation (CV) for all ELISA kits was less than 10.0%. We employed sample dilutions for samples containing biomarkers with elevated concentrations, diluting them 1:5 with the sample diluent provided with the kit.

### Biostatistical analysis

Based on the statistical distribution, the analysis divided the variables into two groups: normally distributed and nonparametric. This was determined by the results of the Kolmogorov-Smirnov test. The results of the normal distribution variables were presented as mean ± SD. The comparison between the patient and control groups was performed using the Mann-Whitney U test for non-normally distributed variables, and one-way analysis of variance for normally distributed scale variables. The contingency tables (χ^2^ tests) assessed the associations between categorical variables. Pearson’s product-moment correlation was used to analyze relationships between scale variables and the biomarkers after transforming non-normally distributed variables into the Ln transformation. Using multivariate general linear model (GLM) analysis (followed by tests of between-subject effects), the associations between categories and biomarkers were investigated, accounting for confounding variables including age, sex, and body mass index (BMI). From the multivariate GLM, we computed the estimated marginal means of variables after controlling all covariates. We performed a binary logistic regression analysis using the diagnosis of severe versus moderate MS and NMS in the PD group, with biomarkers serving as explanatory variables. The odds ratio, along with its 95% confidence intervals, was calculated, as well as the predictive accuracy, sensitivity, and specificity. The latter was used to estimate the model’s effect size. Subsequently, we computed the principal component (PC) scores from the first PC extracted from the scales of overall severity of PD (OSOP), namely, TOTAL-nM-EDL, psychiatric symptoms, TOTAL-m-EDL, TOTAL-PARTIII, TOTAL-Part IV, and Hoehn & Yahr Stage. This PC analysis complies with stringent criteria, including Kaiser-Meyer-Olkin Measure of Sampling Adequacy (KMO) value > 0.8 (actually it is 0.854), a significant Bartlett’s test of sphericity is significant (χ^2^ = 1919.692, df = 15, p < 0.001), explained variance>50% (actually it is 87.20%), and all loadings of the first PC > 0.7 (actually all are higher than 0.921). In this study, statistical significance was established at a p-value of 0.05 using two-tailed tests. IBM SPSS 26 for Windows was utilized to analyze the data. G*Power 3.1.9.7 showed that the a priori estimated sample size was 136, given a power of 0.90, alpha = 0.05, and effect size = 0.28.

## Results

### Characteristics of PD patients and control groups

**Table 1** shows the socio-demographic and clinical characteristics of the PD and healthy control groups. Both groups were comparable in terms of age, sex, body mass index (BMI), and key socio-demographic factors, including tobacco use, occupational status, and marital status. PD patients had significantly higher scores on all clinical assessments of CFS, neurological deficits, motor symptoms, and disease severity (Hoehn & Yahr stage) compared to controls.

**Table 1.** Demographic and clinical data in healthy controls (HC) and patients with Parkinson’s disease.

| **Parameters** | **HC**<br>**n = 45** | **PD**<br>**n = 90** | **df** | **F** | **p** |
| --- | --- | --- | --- | --- | --- |
| Age | 61.3±5.3 | 63.7±13.2 | 1/133 | 1.392 | 0.240 |
| Sex (F/M) | 19/26 | 40/50 | 1 | 0.600 | 0.806 |
| BMI | 27.02±2.27 | 26.80±4.13 | 1/133 | 0.113 | 0.737 |
| TUD (No/Yes) | 31/14 | 72/18 | 1 | 2.048 | 0.152 |
| Employment (No/Yes) | 36/9 | 79/11 | 1 | 1.438 | 0.230 |
| Marital State (Single/Married) | 5/40 | 19/71 | 1 | 2.052 | 0.152 |
| Total FF | 11.6±2.6 | 40.3±7.3 | 1/133 | MWU | <0.001 |
| Pure FF | 8.9±2.3 | 29.2±6.2 | 1/133 | MWU | <0.001 |
| TOTAL-nM-EDL | 0 | 31.07±5.7 | / | MWU | <0.001 |
| Psychiatric symptoms | 0 | 21.8±3.5 | / | MWU | <0.001 |
| TOTAL-m-EDL | 0 | 30.00±6.9 | / | MWU | <0.001 |
| TOTAL-PART III | 0 | 54.3±11.4 | / | MWU | <0.001 |
| TOTAL-Part IV | 0 | 16.8±4.1 | / | MWU | <0.001 |
| Hoehn & Yahr Stage | 0 | 2.8±0.9 | / | MWU | <0.001 |
| Levodopa (No/Yes) | / | 27/63 | / | / | <0.001 |
| Kemadrin (No/Yes) | / | 65/25 | / | / | <0.001 |
| Sinamet (No/Yes) | / | 67/23 | / | / | <0.001 |

BMI: body mass index; TUD: Tobacco use disorder; FF: Fabro-Fatigue; EDL: Experiences of Daily Living; MWU: Mann-Whitney U test

### Concentrations of Serum Biomarkers

An analysis of serum biomarkers revealed significant disparities between the PD and HC groups (**Table 2**). The levels of GFAP, HMGB1, BDNF, NSE, UCTHL1, DKK1, and α-Synuclein were markedly elevated in patients with PD. In contrast, no notable differences were seen between groups for R-spondin 1 (RSPO1), sclerostin, or β-Amyloid-42 levels.

**Table 2.** Estimated marginal mean of biomarkers in PD and healthy control groups.

| **Dependent Variable** | **HC** | **PD** | **df** | **F** | ***p*** |
| --- | --- | --- | --- | --- | --- |
| GFAP (pg/mL) | 1.658(0.206) | 2.485(0.145) | 1/129 | 10.631 | 0.001 |
| HMGB1 (pg/mL) | 2844.913(274.571) | 3924.264(193.209) | 1/129 | 10.203 | 0.002 |
| BDNF (pg/mL) | 20.779(2.751) | 28.342(1.936) | 1/129 | 4.989 | 0.027 |
| RSPO1 (ng/mL) | 207.824(17.761) | 237.101(12.498) | 1/129 | 1.794 | 0.183 |
| NSE (ng/mL) | 1.366(0.171) | 2.73(0.12) | 1/129 | 42.075 | <0.001 |
| UCTHL1 (ng/mL) | 1.59(0.168) | 2.105(0.118) | 1/129 | 6.181 | 0.014 |
| DKK1 (ng/mL) | 14.752(1.863) | 27.619(1.311) | 1/129 | 31.512 | <0.001 |
| Alpha-Synuclein (ng/mL) | 11.305(2.132) | 17.227(1.501) | 1/129 | 5.092 | 0.026 |
| Sclerostin (ng/mL) | 12.855(1.328) | 12.195(0.935) | 1/129 | 0.163 | 0.687 |
| Beta-Amyloid-42 (pg/mL) | 38.132(4.023) | 47.332(2.831) | 1/129 | 3.453 | 0.065 |

GFAP: glial fibrillary acidic protein, HMGB1: high-mobility group box 1, BDNF: brain-derived neurotrophic factor, RSPO1: R-spondin-1; NSE: neuron-specific enolase, UCTHL1: ubiquitin carboxyl-terminal hydrolase L1, and DKK1: Dickkopf-1.

### Biomarkers as Discriminatory Tools for PD Diagnosis

A binary logistic regression model was constructed to identify the most effective markers for distinguishing patients with PD from healthy controls (**Table 3**). The model, including NSE, DKK1, and Beta-Amyloid-42 as explanatory variables, was statistically significant (χ² = 89.046, *p* < 0.001; Nagelkerke R² = 0.671). The Receiver Operating Characteristic (ROC) curve analysis for this biomarker panel demonstrated high diagnostic efficacy, with an Area Under the Curve (AUC) of 0.932 (95% CI: 0.894-0.971, *p* < 0.001). The model demonstrated good accuracy in detecting PD, with 77.8% sensitivity, 85.6% specificity, and an overall accuracy of 83.0%. (Gini Index: 0.864; Kolmogorov-Smirnov (K-S) statistic: 0.733).

**Table 3.** Results of binary logistic regression analyses with Parkinson’s disease (PD) as the dependent variable and biomarkers as explanatory variables.

| **Dichotomies** | **Explanatory variables** | **$\beta$** | **SE** | **Wald** | **df** | ***p*** | **OR** | **95% CI** |
| --- | --- | --- | --- | --- | --- | --- | --- | --- |
| PD vs. controls | NSE | 2.021 | 0.491 | 16.924 | 1 | <0.001 | 7.545 | 2.881-19.76 |
|  | DKK1 | 1.684 | 0.503 | 11.221 | 1 | <0.001 | 5.387 | 2.011-14.429 |
| | β-Amyloid-42 | 0.677 | 0.279 | 5.905 | 1 | 0.015 | 1.968 | 1.140-3.397 |

NSE: neuron-specific enolase and DKK1: Dickkopf-1.

### Associating Biomarkers with Clinical Severity in PD

We used multiple linear regression to examine the relationship between serum biomarker levels and the severity of several clinical domains (**Table 4**). The results consistently indicated NSE, DKK1, and β-Amyloid-42 as significant predictors across several neuropsychiatric and motor assessments. Regression models were established for pure FF, psychiatric symptoms, total-mEDL, total-PART III, and PART IV, with NSE and DKK1 consistently recognized as the principal variables. For example, we found that 49.1% of the variance in overall PD severity was explained by DKK1, NSE, age, and β-Amyloid-42 (all positively associated).

**Table 4.** Results of multiple regression with the scores of the neuropsychiatric scales as dependent variables, and serum biomarkers as explanatory variables in Parkinson’s disease patients.

| Dependent Variables | Explanatory Variables | Coefficient statistics |  |  | Model statistics |  |  |  |
| --- | --- | --- | --- | --- | --- | --- | --- | --- |
| | | $\beta$ | t | p | F model | df | p | R<sup>2</sup> |
| 1. Pure FF | **Model** |  |  |  | 22.91 | 4/130 | <0.001 | 0.413 |
|  | NSE | 0.407 | 5.45 | <0.001 |  |  |  |  |
|  | DKK1 | 0.244 | 3.23 | 0.002 |  |  |  |  |
| | β-Amyloid-42 | 0.182 | 2.66 | 0.009 | | | | |
|  | UCTHL1 | 0.151 | 2.20 | 0.029 |  |  |  |  |
| 2. Psychiatric symptoms | **Model** |  |  |  | 32.87 | 3/131 | <0.001 | 0.429 |
|  | NSE | 0.396 | 5.41 | <0.001 |  |  |  |  |
|  | DKK1 | 0.329 | 4.45 | <0.001 |  |  |  |  |
| | β-Amyloid-42 | 0.181 | 2.70 | 0.008 | | | | |
| 3. TOTAL -mEDL | **Model** |  |  |  | 28.53 | 4/130 | <0.001 | 0.467 |
|  | DKK1 | 0.346 | 4.79 | <0.001 |  |  |  |  |
|  | NSE | 0.364 | 5.13 | <0.001 |  |  |  |  |
|  | Age | 0.221 | 3.43 | <0.001 |  |  |  |  |
| | β-Amyloid-42 | 0.138 | 2.13 | 0.035 | | | | |
| 4. TOTAL-PART III | **Model** |  |  |  | 30.10 | 4/130 | <0.001 | 0.481 |
|  | DKK1 | 0.336 | 4.72 | <0.001 |  |  |  |  |
|  | NSE | 0.364 | 5.20 | <0.001 |  |  |  |  |
|  | Age | 0.242 | 3.80 | <0.001 |  |  |  |  |
|  | β-Amyloid-42 | 0.165 | 2.57 | 0.011 |  |  |  |  |
| 5. TOTAL-Part IV | **Model** |  |  |  | 30.03 | 4/130 | <0.001 | 0.480 |
|  | DKK1 | 0.326 | 4.571 | <0.001 |  |  |  |  |
|  | NSE | 0.357 | 5.096 | <0.001 |  |  |  |  |
|  | Age | 0.277 | 4.350 | <0.001 |  |  |  |  |
|  | β-Amyloid-42 | 0.155 | 2.411 | 0.017 |  |  |  |  |
| 6. Overall severity of PD | **Model** |  |  |  | 31.33 | 4/130 | <0.001 | 0.491 |
|  | DKK1 | 0.350 | 4.95 | <0.001 |  |  |  |  |
|  | NSE | 0.380 | 5.48 | <0.001 |  |  |  |  |
|  | Age | 0.199 | 3.15 | 0.002 |  |  |  |  |
|  | β-Amyloid-42 | 0.171 | 2.68 | 0.008 |  |  |  |  |

NSE: neuron-specific enolase; DKK1: Dickkopf-1; UCTHL1: ubiquitin carboxyl-terminal hydrolase L1

## Discussion

This study investigated a panel of blood biomarkers associated with the Wnt pathway and neuronal injury to evaluate their utility in diagnosing PD and predicting clinical severity, particularly CFS-related fatigue and psychiatric symptoms. Our principal findings indicate that a biomarker combination comprising NSE, DKK1, and β-Amyloid-42 has significant diagnostic value, underscoring the potential contribution of neuronal injury, Wnt signaling dysregulation, and amyloid pathology to PD.

The predictive significance of NSE, a well-established marker of neuronal damage [7, 27], confirms that ongoing neurodegeneration is a notable and quantifiable feature of PD. The release of NSE into the extracellular space reflects substantial neuronal injury [28], and our findings align with studies reporting elevated NSE levels in PD and linking them to the disease pathophysiology and progression [29, 30]. Increased NSE concentrations in CSF have been observed in PD patients compared with healthy controls, and these elevations correlate with disease severity, as measured by the Hoehn-Yahr scale and UPDRS part III, suggesting that NSE may serve as a potential marker of PD progression [30].

The inclusion of β-Amyloid-42 in our diagnostic model, despite the absence of significant difference in its mean levels between groups, suggests that this biomarker may contribute to disease severity rather than group discrimination. This observation supports the concept of shared pathogenic mechanisms between PD and other amyloid-related disorders [14, 31]. It is plausible that β-Amyloid-42 interacts with additional pathological processes, such as α-synuclein aggregation, thereby contributing to disease progression even in peripheral contexts [32, 33].

Multiple linear regression analyses consistently identified NSE and DKK1 as the principal predictors of clinical severity across various domains, including overall PD severity, CFS-related fatigue, psychiatric symptoms, and motor impairment. These findings suggest that neuronal damage, reflected by NSE, and impaired Wnt-mediated synaptic support, indicated by DKK1, may progress in parallel with clinical deterioration. The significant elevation of DKK1 in our PD cohort, along with its substantial independent predictive value, highlights the potential role of Wnt signaling disruption in PD pathology. The Wnt pathway plays a crucial role in neural development and synaptic plasticity [18], and its inhibition by DKK1 may contribute to synaptic dysfunction in PD [6]. Experimental studies further demonstrate that inducible DKK1 expression in the striatum leads to the loss of inhibitory synapses and compromised motor function, underscoring the significance of Wnt signaling in synaptic integrity [34, 35]. Moreover, the interaction between synaptic dysfunction and neuronal injury suggests that both NSE and DKK1 may serve as valuable indicators of PD progression [36]. In experimental PD models, reduced Wnt signaling has also been linked to increased oxidative stress and exacerbated neuronal injury, further supporting a mechanistic connection between Wnt pathway dysfunction and disease progression [37].

It was somewhat unexpected that α-synuclein, a well-established PD marker, showed elevated levels yet was excluded from the final model predicting motor symptom severity. This finding may reflect the complex metabolism of α-synuclein, whereby its aggregation in the CNS may lead to reduced or variable levels in peripheral fluids [38]. Alternatively, specific molecular forms of α-synuclein, such as oligomeric or phosphorylated forms - which were not assessed in the present study – may have greater clinical relevance for disease progression and symptom severity [39, 40].

Our comprehensive analysis also provides context for the additional biomarkers investigated. The observed elevation of HMGB1 and UCHL1 in the PD cohort aligns with prior studies highlighting the roles of neuroinflammation and ubiquitin-proteasome system dysfunction in PD pathophysiology [10, 16]. However, their exclusion from the final predictive models suggests that, within this cohort, their serum levels may have lower discriminative capacity compared with the primary biomarker panel comprising NSE, DKK1, and β-Amyloid-42 for the outcomes assessed.

Previous studies have reported that higher BDNF levels have been associated with more advanced stages of PD, possibly reflecting a compensatory response [41, 42]. However, other studies have observed reduced BDNF levels in PD, highlighting the complex and potentially stage-dependent role of BDNF in disease progression [41, 43]. Furthermore, GFAP demonstrated a notable correlation with motor severity scores (UPDRS PART III & IV), highlighting the impact of neuroinflammation and astrocytic activation on disease progression [44, 45]. Although GFAP levels were elevated in PD patients compared to controls, they were not retained as a primary component of our diagnostic model.

The identification of a robust three-biomarker serum profile for PD diagnosis may have important clinical implications. Recent proteomic and machine learning studies have identified proteins panels capable of predicting PD years before symptom onset, suggesting the feasibility of early blood-based diagnosis approaches [46]. Similarly, CSF biomarkers such as α-synuclein, β-amyloid-42, and tau have shown potential diagnostic value in PD [47]. In our study, a substantial proportion of the variance in chronic fatigue symptoms attributable to Parkinson’s disease was explained by four biomarkers reflecting neuronal damage, disrupted neuronal homeostasis, and Wnt pathway dysregulation. Mechanistically, fatigue may arise from diverse biological stressors that converge on Wnt/β-catenin signaling pathways, thereby disrupting cellular homeostatic [48]. Aberrations in this pathway are implicated in synaptic loss, blood–brain barrier breakdown, and neurodegenerative processes characteristic of Parkinson’s disease [49]. Consistent with this framework, elevated biomarkers of neuronal or astroglial injury have been reported not only in PD [50] but also in chronic fatigue syndrome [51, 52]. Significant correlations between fatigue severity and biomarkers such as neurofilament light chain and GFAP have also been observed in other conditions, including thalassemia [53]. Furthermore, fatigue severity in PD has been directly associated with biomarkers of neuronal damage [54, 55], and a large proportion of variance in fatigue scores could be predicted by inflammatory and neuronal autoantibodies [56]. Collectively, these findings support a model in which neuronal injury – arising from neurodegeneration, systemic inflammation, or autoimmune processes - contributes to the pathogenesis of fatigue across multiple disorders.

## Limitations

This research presents several issues. The cross-sectional design identifies associations but does not establish causal relationships. Longitudinal studies are crucial for validating these biomarkers as indicators of disease progression. Furthermore, while our panel was comprehensive, the diagnostic and prognostic utility of α-synuclein subtypes (oligomers, phosphorylated) in serum necessitates additional exploration, as they may carry additional clinical relevance [40, 57]. Ultimately, future studies in independent cohorts are needed to confirm the generalizability of our findings.

## Conclusion

In conclusion, our findings suggest that a multi-biomarker approach focusing on neuronal damage pathways, Wnt signaling, and amyloid metabolism may represent an effective diagnostic strategy for assessing disease severity. Notably, NSE and DKK1 were identified as key factors influencing clinical severity, including CFS and psychiatric symptoms.

## Ethics approval

The study was granted approval by the Institutional Ethics Committee of the University of Kufa (MEC-110/2025).

## Acknowledgments

We express our profound gratitude to the neurologists Dr. Mohsen Mohammed Musa Al-Najm, a faculty member at the College of Medicine, Jaber Ibn Hayyan University, and Dr. Hussein Abdulkarim Al-Barzanji, a consultant at Al-Najaf Al-Ashraf Teaching Hospital, for their assistance.

## Data availability

The dataset compiled in this study will be made available on request by the last author (MM).

## References

1. Steinmetz JD, Seeher KM, Schiess N, Nichols E, Cao B, Servili C, Cavallera V, Cousin E, Hagins H, Moberg ME: Global, regional, and national burden of disorders affecting the nervous system, 1990–2021: a systematic analysis for the Global Burden of Disease Study 2021. The Lancet Neurology 2024, 23(4):344–381.

2. Schapira AH, Chaudhuri KR, Jenner P: Non-motor features of Parkinson disease. Nature Reviews Neuroscience 2017, 18(7):435–450.

3. Weintraub D, Mamikonyan E: The neuropsychiatry of Parkinson disease: a perfect storm. The American Journal of Geriatric Psychiatry 2019, 27(9):998–1018.

4. Memou A, Dimitrakopoulos L, Kedariti M, Kentros M, Lamprou A, Petropoulou-Vathi L, Valkimadi P-E, Rideout HJ: Defining (and blocking) neuronal death in Parkinson’s disease: Does it matter what we call it? Brain Research 2021, 1771:147639.

5. Benskey MJ, Sellnow RC, Sandoval IM, Sortwell CE, Lipton JW, Manfredsson FP: Silencing alpha synuclein in mature nigral neurons results in rapid neuroinflammation and subsequent toxicity. Frontiers in Molecular Neuroscience 2018, 11:36.

6. Calabresi P, Mechelli A, Natale G, Volpicelli-Daley L, Di Lazzaro G, Ghiglieri V: Alpha-synuclein in Parkinson’s disease and other synucleinopathies: from overt neurodegeneration back to early synaptic dysfunction. Cell death & disease 2023, 14(3):176.

7. Dutta R: Role of neuron specific enolase as a biomarker in Parkinson’s disease. J Neurosci Neurol Disorders 2021, 5:061–068.

8. Dichev V, Kazakova M, Sarafian V: YKL-40 and neuron-specific enolase in neurodegeneration and neuroinflammation. Reviews in the Neurosciences 2020, 31(5):539–553.

9. Liu Z, Meray RK, Grammatopoulos TN, Fredenburg RA, Cookson MR, Liu Y, Logan T, Lansbury Jr PT: Membrane-associated farnesylated UCH-L1 promotes α-synuclein neurotoxicity and is a therapeutic target for Parkinson’s disease. Proceedings of the National Academy of Sciences 2009, 106(12):4635–4640.

10. Buneeva O, Medvedev A: Ubiquitin Carboxyl-Terminal Hydrolase L1 and Its Role in Parkinson’s Disease. In: International Journal of Molecular Sciences. vol. 25; 2024.

11. Minami A, Nakanishi A, Matsuda S, Kitagishi Y, Ogura Y: Function of α-synuclein and PINK1 in Lewy body dementia (Review). Int J Mol Med 2015, 35(1):3–9.

12. Shu H, Zhang P, Gu L: Alpha-synuclein in peripheral body fluid as a biomarker for Parkinson’s disease. Acta Neurologica Belgica 2024, 124(3):831–842.

13. Siderowf A, Xie S, Hurtig H, Weintraub D, Duda J, Chen-Plotkin A, Shaw L, Van Deerlin V, Trojanowski J, Clark C: CSF amyloid β 1-42 predicts cognitive decline in Parkinson disease. Neurology 2010, 75(12):1055–1061.

14. Xiang C, Cong S, Tan X, Ma S, Liu Y, Wang H, Cong S: A meta-analysis of the diagnostic utility of biomarkers in cerebrospinal fluid in Parkinson’s disease. npj Parkinson’s Disease 2022, 8(1):165.

15. Nikitina MA, Koroleva ES, Brazovskaya NG, Boyko AS, Levchuk LA, Ivanova SA, Alifirova VM: Associations of serum neuromarkers with clinical features of Parkinson’s disease. Zhurnal Nevrologii i Psikhiatrii Imeni SS Korsakova 2024, 124(4):145–152.

16. Ma Z-l, Wang Z-l, Zhang F-y, Liu H-x, Mao L-h, Yuan L: Biomarkers of Parkinson’s disease: from basic research to clinical practice. Aging and Disease 2024, 15(4):1813.

17. Gray LKC: Understanding the role of astrocytes in a rare model of Parkinson’s disease. University of Otago; 2024.

18. Gopar-Cuevas Y, Duarte-Jurado AP, Diaz-Perez RN, Saucedo-Cardenas O, Loera-Arias MJ, Montes-de-Oca-Luna R, Rodriguez-Rocha H, Garcia-Garcia A: Pursuing multiple biomarkers for early idiopathic Parkinson’s disease diagnosis. Molecular neurobiology 2021, 58(11):5517–5532.

19. Postuma RB, Poewe W, Litvan I, Lewis S, Lang AE, Halliday G, Goetz CG, Chan P, Slow E, Seppi K et al: Validation of the MDS clinical diagnostic criteria for Parkinson’s disease. Movement Disorders 2018, 33(10):1601–1608.

20. Horváth K, Aschermann Z, Ács P, Deli G, Janszky J, Komoly S, Balázs É, Takács K, Karádi K, Kovács N: Minimal clinically important difference on the Motor Examination part of MDS-UPDRS. Parkinsonism & related disorders 2015, 21(12):1421–1426.

21. Storch A, Schneider CB, Klingelhöfer L, Odin P, Fuchs G, Jost WH, Martinez-Martin P, Koch R, Reichmann H, Chaudhuri KR: Quantitative assessment of non-motor fluctuations in Parkinson’s disease using the Non-Motor Symptoms Scale (NMSS). Journal of Neural Transmission 2015, 122(12):1673–1684.

22. Rabey JM, Korczyn AD: The Hoehn and Yahr rating scale for Parkinson’s disease. In: Instrumental methods and scoring in extrapyramidal disorders. Springer; 1995: 7–17.

23. Al-Hakeim HK, Al-Rammahi DA, Al-Dujaili AH: IL-6, IL-18, sIL-2R, and TNFα proinflammatory markers in depression and schizophrenia patients who are free of overt inflammation. Journal of affective disorders 2015, 182:106–114.

24. Goetz CG, Fahn S, Martinez-Martin P, Poewe W, Sampaio C, Stebbins GT, Stern MB, Tilley BC, Dodel R, Dubois B: Movement Disorder Society-sponsored revision of the Unified Parkinson’s Disease Rating Scale (MDS-UPDRS): process, format, and clinimetric testing plan. Movement disorders 2007, 22(1):41–47.

25. Goetz CG, Tilley BC, Shaftman SR, Stebbins GT, Fahn S, Martinez-Martin P, Poewe W, Sampaio C, Stern MB, Dodel R: Movement Disorder Society-sponsored revision of the Unified Parkinson’s Disease Rating Scale (MDS-UPDRS): scale presentation and clinimetric testing results. Movement disorders: official journal of the Movement Disorder Society 2008, 23(15):2129–2170.

26. Zachrisson O, Regland B, Jahreskog M, Kron M, Gottfries CG: A rating scale for fibromyalgia and chronic fatigue syndrome (the FibroFatigue scale). Journal of psychosomatic research 2002, 52(6):501–509.

27. Chaves ML, Camozzato AL, Ferreira ED, Piazenski I, Kochhann R, Dall’Igna O, Mazzini GS, Souza DO, Portela LV: Serum levels of S100B and NSE proteins in Alzheimer’s disease patients. Journal of neuroinflammation 2010, 7:6.

28. Constantinescu R, Zetterberg H, Holmberg B, Rosengren L: Levels of brain related proteins in cerebrospinal fluid: an aid in the differential diagnosis of parkinsonian disorders. Parkinsonism Relat Disord 2009, 15(3):205–212.

29. Hou C, Yang F, Li S, Ma HY, Li FX, Zhang W, He W: A nomogram based on neuron-specific enolase and substantia nigra hyperechogenicity for identifying cognitive impairment in Parkinson’s disease. Quant Imaging Med Surg 2024, 14(5):3581–3592.

30. Papuć E, Rejdak K: Increased Cerebrospinal Fluid S100B and NSE Reflect Neuronal and Glial Damage in Parkinson’s Disease. Front Aging Neurosci 2020, 12:156.

31. Riboldi GM, Vialle RA, Navarro E, Udine E, de Paiva Lopes K, Humphrey J, Allan A, Parks M, Henderson B, Astudillo K: Transcriptome deregulation of peripheral monocytes and whole blood in GBA-related Parkinson’s disease. Molecular Neurodegeneration 2022, 17(1):52.

32. Saitgareeva AR, Bulygin KV, Gareev IF, Beylerli OA, Akhmadeeva LR: The role of microglia in the development of neurodegeneration. Neurological Sciences 2020, 41(12):3609–3615.

33. Masliah E, Rockenstein E, Veinbergs I, Sagara Y, Mallory M, Hashimoto M, Mucke L: beta-amyloid peptides enhance alpha-synuclein accumulation and neuronal deficits in a transgenic mouse model linking Alzheimer’s disease and Parkinson’s disease. Proc Natl Acad Sci U S A 2001, 98(21):12245–12250.

34. Galli S, Stancheva SH, Dufor T, Gibb AJ, Salinas PC: Striatal synapse degeneration and dysfunction are reversed by reactivation of wnt signaling. Frontiers in synaptic neuroscience 2021, 13:670467.

35. Galli S, Lopes DM, Ammari R, Kopra J, Millar SE, Gibb A, Salinas PC: Deficient Wnt signalling triggers striatal synaptic degeneration and impaired motor behaviour in adult mice. Nature communications 2014, 5(1):4992.

36. Nilsson J, Constantinescu J, Nellgård B, Jakobsson P, Brum WS, Gobom J, Forsgren L, Dalla K, Constantinescu R, Zetterberg H: Cerebrospinal fluid biomarkers of synaptic dysfunction are altered in Parkinson’s disease and related disorders. Movement Disorders 2023, 38(2):267–277.

37. Stephano F, Nolte S, Hoffmann J, El-Kholy S, Von Frieling J, Bruchhaus I, Fink C, Roeder T: Impaired Wnt signaling in dopamine containing neurons is associated with pathogenesis in a rotenone triggered Drosophila Parkinson’s disease model. Scientific reports 2018, 8(1):2372.

38. Mollenhauer B, Caspell-Garcia CJ, Coffey CS, Taylor P, Singleton A, Shaw LM, Trojanowski JQ, Frasier M, Simuni T, Iranzo A et al: Longitudinal analyses of cerebrospinal fluid α-Synuclein in prodromal and early Parkinson’s disease. Movement disorders : official journal of the Movement Disorder Society 2019, 34(9):1354–1364.

39. Li XY, Li W, Li X, Li XR, Sun L, Yang W, Cai Y, Chen Z, Wu J, Wang C et al: Alterations of Erythrocytic Phosphorylated Alpha-Synuclein in Different Subtypes and Stages of Parkinson’s Disease. Front Aging Neurosci 2021, 13:623977.

40. Chen WR, Chen JC, Chang SY, Chao CT, Wu YR, Chen CM, Chou C: Phosphorylated α-synuclein in diluted human serum as a biomarker for Parkinson’s disease. Biomedical journal 2022, 45(6):914–922.

41. Scalzo P, Kümmer A, Bretas TL, Cardoso F, Teixeira AL: Serum levels of brain-derived neurotrophic factor correlate with motor impairment in Parkinson’s disease. J Neurol 2010, 257(4):540–545.

42. Huang Y, Huang C, Yun W: Peripheral BDNF/TrkB protein expression is decreased in Parkinson’s disease but not in Essential tremor. Journal of clinical neuroscience : official journal of the Neurosurgical Society of Australasia 2019, 63:176–181.

43. Chen Z, Zhang H: A meta-analysis on the role of brain-derived neurotrophic factor in Parkinson’s disease patients. Advances in Clinical and Experimental Medicine 2023, 32(3):285–295.

44. Lin J, Ou R, Li C, Hou Y, Zhang L, Wei Q, Pang D, Liu K, Jiang Q, Yang T et al: Plasma glial fibrillary acidic protein as a biomarker of disease progression in Parkinson’s disease: a prospective cohort study. BMC Med 2023, 21(1):420.

45. Tang Y, Han L, Li S, Hu T, Xu Z, Fan Y, Liang X, Yu H, Wu J, Wang J: Plasma GFAP in Parkinson’s disease with cognitive impairment and its potential to predict conversion to dementia. NPJ Parkinsons Dis 2023, 9(1):23.

46. Hällqvist J, Bartl M, Dakna M, Schade S, Garagnani P, Bacalini M-G, Pirazzini C, Bhatia K, Schreglmann S, Xylaki M: Plasma proteomics identify biomarkers predicting Parkinson’s disease up to 7 years before symptom onset. Nature communications 2024, 15(1):4759.

47. Kruse N, Schlossmacher MG, Schulz-Schaeffer WJ, Vanmechelen E, Vanderstichele H, El-Agnaf OM, Mollenhauer B: A first tetraplex assay for the simultaneous quantification of total α-synuclein, tau, β-amyloid42 and dj-1 in human cerebrospinal fluid. Plos one 2016, 11(4):e0153564.

48. Maes M, Kubera M, Kotańska M: Aberrations in the Cross-Talks Among Redox, Nuclear Factor-κB, and Wnt/β-Catenin Pathway Signaling Underpin Myalgic Encephalomyelitis and Chronic Fatigue Syndrome. Frontiers in psychiatry 2022, 13:822382.

49. Marchetti B: Wnt/β-Catenin Signaling Pathway Governs a Full Program for Dopaminergic Neuron Survival, Neurorescue and Regeneration in the MPTP Mouse Model of Parkinson’s Disease. Int J Mol Sci 2018, 19(12).

50. Andersson M, Zetterberg H, Minthon L, Blennow K, Londos E: The cognitive profile and CSF biomarkers in dementia with Lewy bodies and Parkinson’s disease dementia. International journal of geriatric psychiatry 2011, 26(1):100–105.

51. Che N, Huang J, Wang S, Jiang Q, Yang T, Xiao Y, Lin J, Fu J, Ou R, Li C: The potential use of plasma NfL as a diagnostic and prognostic biomarker of fatigue in early Parkinson’s disease. Therapeutic Advances in Neurological Disorders 2025, 18:17562864251324406.

52. Bark L, Larsson I-M, Wallin E, Simrén J, Zetterberg H, Lipcsey M, Frithiof R, Rostami E, Hultström M: Central nervous system biomarkers GFAp and NfL associate with post-acute cognitive impairment and fatigue following critical COVID-19. Scientific reports 2023, 13(1):13144.

53. Ridhaa MAS, Al-Hakeim HK, Kahlol MK, Al-Naqeeb TH, Niu M, Maes M: In transfusion-dependent thalassemia, neuronal damage biomarkers are associated with affective and chronic fatigue symptoms. Sci Rep 2025, 15(1):32721.

54. Wang H, Liu Y, Zhao J, Guo X, Hu M, Chen Y: Possible inflammatory mechanisms and predictors of Parkinson’s disease patients with fatigue (brief review). Clinical Neurology and Neurosurgery 2021, 208:106844.

55. De Dreu MJ, Schouwenaars IT, Rutten GJM, Ramsey NF, Jansma JM: Fatigue in brain tumor patients, towards a neuronal biomarker. NeuroImage: Clinical 2020, 28:102406.

56. Almulla AF, Maes M, Zhou B, Al-Hakeim HK, Vojdani A: Brain-targeted autoimmunity is strongly associated with Long COVID and its chronic fatigue syndrome as well as its affective symptoms. Journal of advanced research 2025, 75:621–633.

57. Zheng H, Xie Z, Zhang X, Mao J, Wang M, Wei S, Fu Y, Zheng H, He Y, Chen H et al: Investigation of α-Synuclein Species in Plasma Exosomes and the Oligomeric and Phosphorylated α-Synuclein as Potential Peripheral Biomarker of Parkinson’s Disease. Neuroscience 2021, 469:79–90.

